# Accuracy and Glycemic Efficacy of Continuous Glucose Monitors in Critically Ill COVID-19 Patients: a Retrospective Study

**DOI:** 10.1101/2022.05.06.22274685

**Authors:** Schafer Boeder, Emily Kobayashi, Gautam Ramesh, Brittany Serences, Kristen Kulasa, Amit R. Majithia

## Abstract

**Background:** Continuous Glucose Monitoring (CGM) is approved for insulin dosing decisions in the ambulatory setting, but not currently for inpatients. CGM has the capacity to reduce patient-provider contact in inpatients with coronavirus disease 2019 (COVID-19), thus potentially reducing in hospital virus transmission. However, there are sparse data on the accuracy and efficacy of CGM to titrate insulin doses in inpatients.

**Methods:** Under an emergency use protocol, CGM (Dexcom G6) was used alongside standard point-of-care (POC) glucose measurements in patients critically ill from complications of COVID-19 requiring intravenous (IV) insulin. Glycemic control during IV insulin therapy was retrospectively assessed comparing periods with and without adjunctive CGM use. Accuracy metrics were computed and Clarke Error Grid analysis performed comparing CGM glucose values with POC measurements.

**Results:** 24 critically ill patients who met criteria for emergency use of CGM resulted in 47333 CGM and 5677 POC glucose values. During IV insulin therapy, individuals’ glycemic control improved when CGM was used (mean difference -30.2 mg/dL). Among 2194 matched CGM:POC glucose pairs a high degree of concordance was observed with a MARD of 14.8% and 99.5% of CGM:POC pairs falling in Zones A and B of the Clarke Error Grid.

**Conclusions:** CGM use in critically ill COVID-19 patients improved glycemic control during IV insulin therapy. CGM glucose data were highly concordant with POC glucose during IV insulin therapy in critically ill patients suggesting that CGM could substitute for POC measurements in inpatients thus reducing patient-provider contact and mitigating infection transmission.

## Introduction

Uncontrolled hyperglycemia is associated with increased length of hospital stay and mortality.[1] Accurate blood glucose measurements are essential for safe and effective titration of insulin particularly in critically ill patients on IV insulin to achieve optimal blood glucose targets.[2] The current standard of care for measuring inpatient blood glucose for insulin dosing decision is to use point-of-care (POC) glucose meter devices which require hospital staff to manually sample patients at frequencies ranging from once every 4-6 hours for those receiving subcutaneous insulin injections, to once every 0.5 to 2 hours for those on intravenous insulin.[2] While POC is the standard of care, clinically significant glycemic events can be missed between POC tests even at the highest sampling frequency of 0.5 hours. Simply increasing the frequency of POC testing would increase burden on the hospital staff who carry out the tests and increase their exposure to COVID-19 and other transmissible infections.

Continuous glucose monitoring (CGM), which requires one sensor insertion every 10-14 days and can sample interstitial glucose every 5 minutes, has the potential to address the shortcomings of POC testing.[3] While non-adjunctive CGM has been approved by the U.S. Food and Drug Administration (FDA) for use in the ambulatory setting since 2016,[4,5] their use in hospitals has not been approved and remains experimental despite mounting evidence that CGM improves glycemic control in inpatients.[6–10] The COVID-19 pandemic accelerated the adoption of CGM technology within hospitals,[11,12] reflecting the need to reduce healthcare provider exposure to severe acute respiratory syndrome coronavirus 2 (SARS-CoV-2) while improving glycemic control in patients with diabetes who are much more likely to be hospitalized after COVID-19 infection than their counterparts without diabetes.[13,14] In April of 2020 the FDA announced its permission for expanded utilization of remote monitoring devices, including the use of CGM in hospitals.[15] More recently, the FDA provided the Dexcom G6 with a breakthrough device designation.

Following the original FDA announcement, our institution developed an emergency protocol for the use of CGM in the ICU to assist with glycemic control in patients with COVID-19 related critical illness requiring intravenous insulin. Here, we report on data collected from 24 individuals relating glycemic outcomes and CGM accuracy to POC measurements.

## Methods

CGM sensors (Dexcom G6, Dexcom, San Diego, CA) were placed on hospitalized patients under an Emergency Operational Need Protocol approved by UC San Diego Health in 2020 allowing use in critically ill patients with COVID-19 requiring continuous intravenous insulin infusion for glycemic control. Under this protocol, a trained physician, nurse practitioner, physician assistant, or diabetes nurse specialist placed the CGM sensor on the abdomen or posterior upper arm of the patient and initiated the system. The CGM receiver (smartphone) was placed within 20 feet of the patient and data were displayed in real time on an iPad (iPad 5, iPadOS 14.7.1, Apple, Cupertino, CA) and monitored by trained nursing staff.

Retrospective analysis of data obtained from the Emergency Protocol was approved by the UC San Diego Institutional Review Board. Data were collected from 23 patients who wore CGM in the hospital between July 2020 and February 2021 and met the following criteria: 1) age ≥18 years, 2) admitted with confirmed COVID-19 infection to an intensive care unit at either UC San Diego Medical Center in Hillcrest or UC San Diego Jacobs Medical Center, and 3) required continuous intravenous insulin infusion therapy. One additional patient did not receive insulin infusion (IV insulin was planned but never implemented) but met all other inclusion criteria. The comparison of glycemic control during insulin infusion achieved with and without CGM excluded data from this patient, but the analysis of CGM accuracy included this patient. CGM data were downloaded using Dexcom CLARITY software. POC glucose data and patient information, including demographics and medical history, were extracted from the electronic health record (EHR) system. Non-numeric glucose value entries were filtered and POC data points with glucose values of less than 20mg/dL were removed as they were considered artifacts.

Data analysis and plotting were implemented in Python 3.9.7 using the standard library, NumPy 1.20.3, SciPy 1.7.1, pandas 1.3.4, Matplotlib 3.4.3 and python-dateutil 2.8.2 packages. Glycemic control during periods ‘on’ and ‘off’ CGM was assessed by comparing POC glucose values during periods of IV insulin infusion with or without CGM use. A Kolmogorov-Smirnov (KS) test and a two-sided student t-test were used to determine statistical significance. CGM and POC pairs were matched based on timepoints, where each POC datapoint was matched with a CGM datapoint within 5 minutes. Clarke Error Grid analysis was computed using a publicly available Python script (https://github.com/suetAndTie/ClarkeErrorGrid) modified to allow for shading zones in different colors. Mean absolute relative difference (MARD) was calculated as previously described.[16]

## Results

Characteristics of the patients included in the study (n=24) are shown in Table 1. Participants were predominantly male (75%) with poorly controlled diabetes (mean A1c 9.8%) and diagnosed with COVID-19 pneumonia/ respiratory failure. Almost all participants required ventilatory (n=23) and hemodynamic (n=22) support with an eventual mortality rate of 54%. For each participant, periods of IV insulin therapy were extracted from the EHR and intersected with CGM and POC glucose values. Figure 1A demonstrates the POC glucose values during insulin infusion – with and without CGM use – for a representative patient.

**Table 1:** Demographics table. Abbreviations: BMI - Body Mass Index, PNA - Pneumonia, ARDS - Acute Respiratory Distress Syndrome

| Patient Demographics | n | % or range |
| --- | --- | --- |
| **Gender** |  |  |
| Male | 18 | 75% |
| Female | 6 | 25% |
| **Anthropometric measurements and lab values** |  |  |
| Age (median) | 61 | 50-84 |
| BMI (median) | 31.0 | 20.5-45.0 |
| A1c on Admission (median) | 8.9% | 6.2%-15.8% |
| **Race/Ethnicity** |  |  |
| Hispanic (non-White) | 11 | 46% |
| White | 9 | 38% |
| Black or African American | 2 | 8% |
| Other, Mixed or Unknown Race | 2 | 8% |
| **Interventions and clinical status** |  |  |
| Mortality | 13 | 54% |
| COVID-19 PNA/ARDS | 24 | 100% |
| Intubation | 23 | 96% |
| Vasopressor | 22 | 92% |
| Glucocorticoids | 22 | 92% |

**Figure 1:**
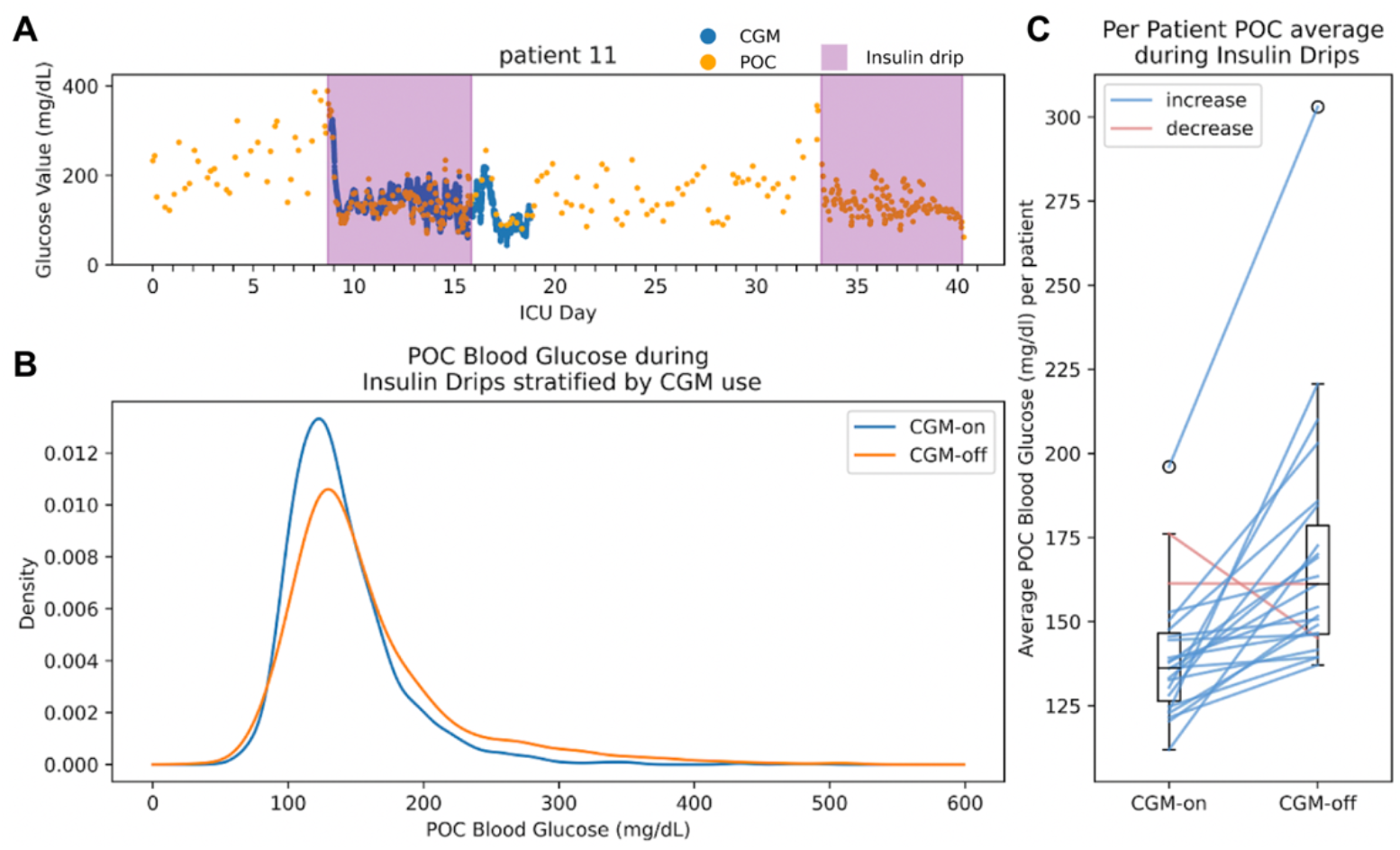
**A)** CGM and POC glucose values plotted by time for a representative patient. Insulin infusion times are highlighted. **B)** Density plot of POC blood glucose values during insulin infusion over all patients. POC values are stratified by concurrent CGM use (n=2101) or no CGM use (n=1739). **C)** Boxplot of average POC values for each patient (n=23) during insulin infusion when on CGM compared to off CGM. Lines connect the means of individual patients. Blue lines indicate an increase in average blood glucose when off CGM. Red lines denote a decrease in average blood glucose when off CGM.

Stratifying the data by CGM use (i.e. ‘CGM-on’ and ‘CGM-off’), 2101 POC glucose measurements coincided with CGM-on and 1739 POC values coincided with CGM-off. The distributions of POC blood glucose levels stratified by CGM use are shown in Figure 1B, demonstrating a decrease in mean glucose (−18.5mg/dL CGM-on vs CGM-off) and fewer extremely high glucose values in the CGM-on distribution. Statistically, these distributions were different under a two sample KS test with test statistic 0.12 and p-value 6.98e-13. We also performed paired analysis to evaluate intraindividual CGM-on and CGM-off periods (Figure 1C), finding a similarly decreased mean POC glucose during CGM-on versus CGM-off (mean decrease -30.2 mg/dL, p<.002 paired t-test). At the individual level, average POC blood glucose during CGM-on versus CGM-off periods decreased in all but two participants.

To evaluate concordance between CGM and POC glucose, POC values were matched to the closest CGM value less than 5 minutes apart. From our dataset (n= 47333 CGM values and n=5677 POC values) 2194 matched CGM:POC pairs were identified. The MARD across all matched pairs was 14.8% ± 0.5. The median ARD was 12.7% (IQR: 6.4%, 20.9%). MARD in the target range of 70-180 mg/dL was 14.9% (n=1811).

To quantify clinical accuracy of CGM in this setting, we performed Clarke Error Grid analysis with the matched CGM:POC data pairs (Figure 2). Values over 400mg/dL were excluded from analysis (2 data points removed) as this exceeds the upper limit of measurement of the Dexcom G6. Of the remaining 2192 matched pairs 73.15% fell into Zone A (clinically accurate), 26.44% fell into Zone B (benign errors that would not lead to inappropriate treatment), 0.14% fell into Zone C (overcorrection errors, harmless corrections), and 0.18% fell into Zone D (dangerous failure to detect hypo- or hyperglycemia). There were no data points in zone E (erroneous treatment error). In summary, 99.5% of CGM:POC pairs fell into Zones A and B of the Clarke Error Grid indicating a high level of clinical accuracy for CGM in critically ill patients on IV insulin.

**Figure 2:**
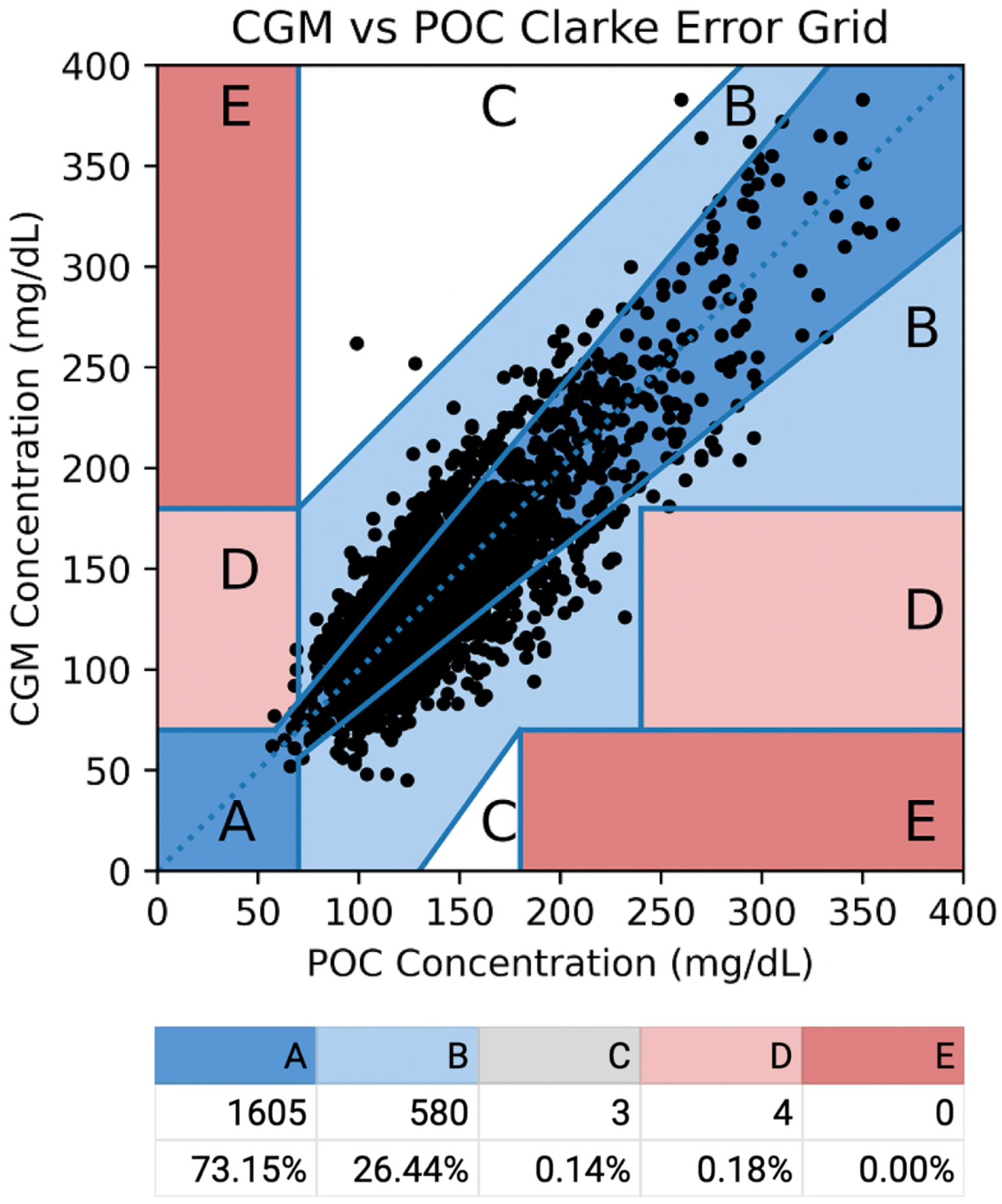
Clarke Error Grid of CGM vs POC blood glucose values in patients with COVID-19 (n=24). Dashed line indicates the 45 degree line. Data is rounded and may not add up to 100%. Two data points fell outside the range of this graph (POC glucose above 400mg/dL) but would have fallen in the A or B zones.

## Discussion

In this retrospective study of adjunctive CGM use in critically ill COVID-19 patients on IV insulin, we found a strong improvement in glycemic control (mean decrease 30.2 mg/dL) during periods of CGM use. An almost perfect clinical concordance was observed between CGM glucose values and POC glucose meters (99.5% CGM:POC matched values falling in Zones A and B of the Clarke Error Grid) suggesting that CGM could safely and effectively substitute POC glucose meters for IV insulin titration in this population where minimizing patient:provider contacts is imperative to infection control.

A major strength of our study includes enrollment of participants with COVID-19 related critical illness. Almost all participants required ventilatory support and over half required hemodynamic support with vasopressors – conditions that could theoretically impact interstitial glucose levels and decrease the concordance between CGM and blood glucose. These factors suggest that the observed concordance of CGM and POC glucose meter values (99.5% Clarke Error Grid zones A and B) is likely a conservative estimate and in the general, non-critically ill inpatient population clinical concordance is likely to be even higher. Furthermore, the improved glycemic control observed during CGM-on versus CGM-off times during intravenous insulin was observed in almost all individual participants (21/23). Taken together, these findings support the use of CGM in critically ill COVID-19 patients in place of POC glucose meters, a timely finding given the trend towards increased infectiousness of newly arising SARS-COV2 variants.[17,18]

Our study has several limitations including a retrospective study design, a relatively small sample size, and lack of a CGM only group. Since patients were not prospectively enrolled, a cryptic bias in the individuals enrolled under the emergency use protocol could confound our findings. Despite a relatively small number of individuals enrolled (n=24), our study is among the largest testing CGM use in critically ill patients to date [7,11,19–23] and the amount of analyzable CGM and POC glucose data obtained (n=47333 CGM values, n=5677 POC values) drives statistically robust inferences. The lack of a CGM only group prevents us from being able to formally test non-adjunctive CGM based insulin titration, but our study evidentiates the safety and potential efficacy of a CGM based insulin titration that needs prospective validation.

## Conclusion

Continuous glucose monitors have not been widely studied in the ICU and have not been approved for inpatient use. In our study of critically ill COVID-19 positive patients on IV insulin, we observe improved glycemic control with adjunctive CGM use compared to standard point of care testing alone. This finding was driven by a reduction in hyperglycemia, which translated to an improvement in average blood glucose during CGM use in 91% of participants. Additionally, CGM demonstrated high concordance with POC, suggesting that it can substitute for POC glucose measurements during IV insulin titration. CGM use would reduce patient provider contact, thereby reducing in hospital transmission of infectious illness such as SARS-Cov2.

## Data Availability

All data produced in the present study are available upon reasonable request to the authors

